# Epidemiological characteristics of the B.1.526 SARS-CoV-2 variant

**DOI:** 10.1101/2021.08.04.21261596

**Authors:** Wan Yang, Sharon K. Greene, Eric R. Peterson, Wenhui Li, Robert Mathes, Laura Graf, Ramona Lall, Scott Hughes, Jade Wang, Anne Fine

## Abstract

To characterize the epidemiological properties of the B.1.526 SARS-CoV-2 variant of interest, here we utilized nine epidemiological and population datasets and model-inference methods to reconstruct SARS-CoV-2 transmission dynamics in New York City, where B.1.526 emerged. We estimated that B.1.526 had a moderate increase (15-25%) in transmissibility and could escape immunity in 0-10% of previously infected individuals. In addition, B.1.526 substantially increased the infection-fatality risk (IFR) among adults 65 or older by >60% during Nov 2020 – Apr 2021, compared to baseline risk estimated for preexisting variants. Overall, findings suggest that new variants like B.1.526 likely spread in the population weeks prior to detection and that partial immune escape (e.g., resistance to therapeutic antibodies) could offset prior medical advances and increase IFR. Early preparedness for and close monitoring of SARS-CoV-2 variants, their epidemiological characteristics, and disease severity are thus crucial to COVID-19 response as it remains a global public health threat.

## INTRODUCTION

The SARS-CoV-2 virus spread quickly worldwide in early 2020, causing the COVID-19 pandemic. As the virus spread, it also diversified, and multiple novel SARS-CoV-2 variants emerged in different populations, producing both local and global waves of infection. Several variants have been characterized as variants of concern (VOC) or of interest (VOI), based on evidence regarding their ability to increase transmissibility, evade immunity conferred by either prior infection or vaccination, or cause more severe disease. Accurately estimating the epidemiological characteristics and impact of these variants is thus important for informing public health response, such as monitoring effectiveness of vaccines and therapeutic antibodies. More broadly, such findings can also provide insights into the long-term trajectory of SARS-CoV-2 beyond the pandemic phase.

The B.1.526 variant (WHO designation: Iota)(*1*), a SARS-CoV-2 VOI, was identified during Nov 2020 and quickly became a predominant variant in the New York City (NYC) area (*2-4*). It has also been detected in all 52 states/territories in the US and at least 27 other countries (GISAID data (*5*), as of 6/9/2021). An initial laboratory study (*2*) suggested that this variant is to some extent resistant to two therapeutic monoclonal antibodies in clinical use and neutralization by convalescent plasma and vaccinee sera. However, another study (*4*) examined all sequenced B.1.526 cases in NYC identified as of April 5, 2021 (n = 3,679) and showed preliminary evidence that this variant did not increase risk for infection after vaccination or reinfection. Both studies may be limited due to the small number of specimens available for analysis as well as delay in observation and reporting. Given these discrepancies, here we utilize detailed population epidemiological data collected since the beginning of the COVID-19 pandemic in NYC (March 1, 2020 – April 30, 2021) and multiple model-inference methods to estimate the transmissibility, immune escape potential, and disease severity of B.1.526. Of note, we include the combination of B.1.526-S:E484K and B.1.526-S:S477N as B.1.526.

As shown in Fig 1 (overall study design), we first apply a network model-inference system to reconstruct underlying SARS-CoV-2 transmission dynamics in NYC, accounting for under-detection of infection. This analysis allows estimation of key population variables and parameters (e.g., the infection rate including those not detected as cases and transmission rate) at the neighborhood-level as well as citywide. As such, we are able to examine in detail the epidemiological dynamics in Washington Heights – Inwood (WHts), the neighborhood where B.1.526 was initially identified (*2*). Using these estimates and additional variant prevalence data, we further apply a city-level multi-variant, age-structured model to estimate the changes in transmissibility and immune escape potential for B.1.526. Lastly, we utilize findings from the first two model systems to estimate variant-specific infection fatality risk (IFR, i.e. the fraction of all persons with SARS-CoV-2 infection who died from the disease), for B.1.526 and B.1.1.7, separately. Overall, our study documents the emergence and impact of B.1.526. We close with a discussion on lessons learned from this SARS-CoV-2 VOI and implications for future COVID-19 pandemic response.

**Fig 1.**
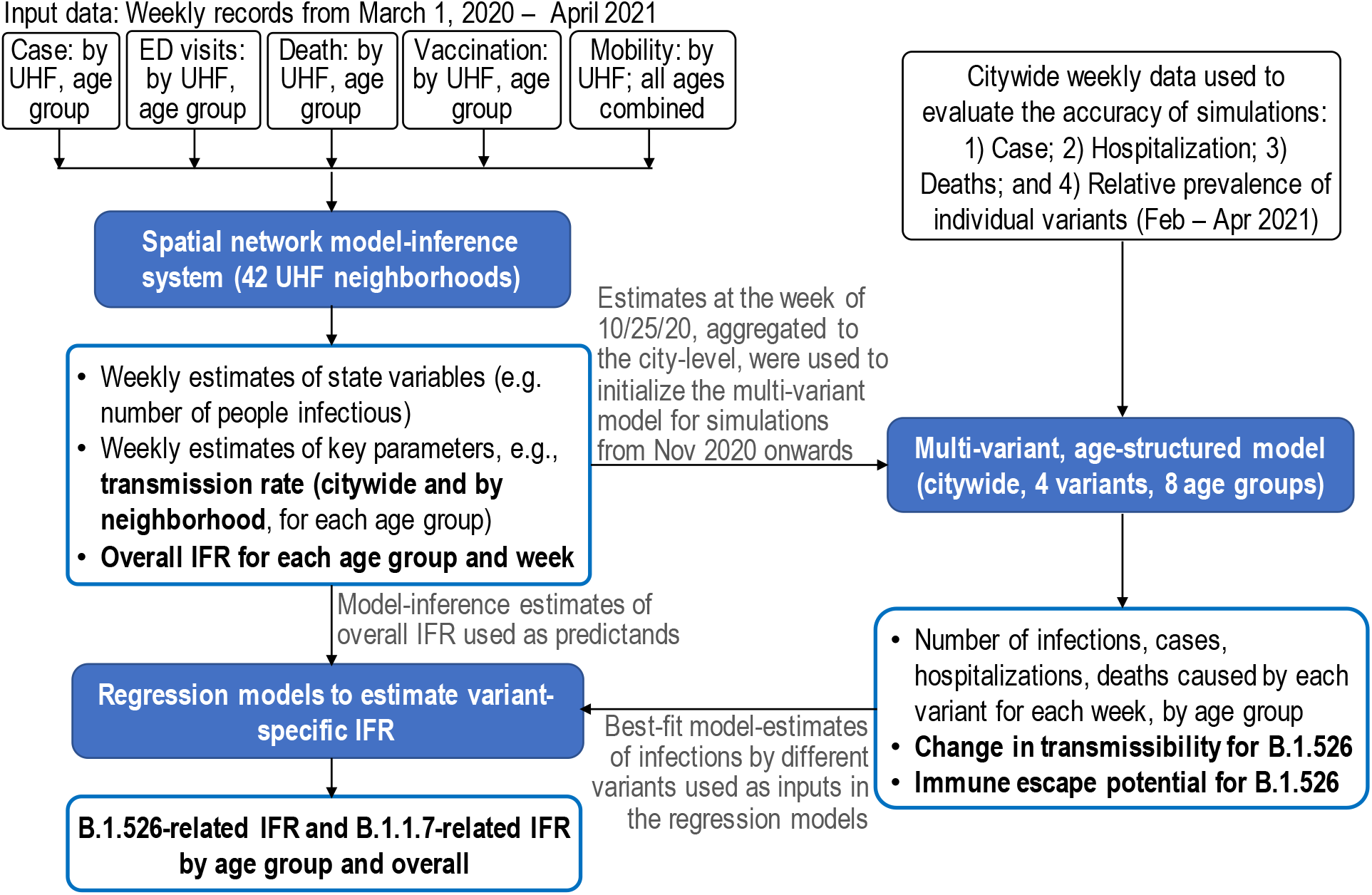
Study design. This study included three modeling analyses: 1) spatial network model- inference to construct the transmission dynamics and estimate key population variables and parameters by United Hospital Fund (UHF) neighborhood of residence and age group; 2) city- level multi-variant, age-structured modeling to simulate and estimate the changes in transmissibility and immune escape potential for B.1.526; and 3) linear regression models to estimate variant-specific infection fatality risk (IFR), for B.1.526 and B.1.1.7, separately. Nine datasets (listed in the black open boxes) were used as model inputs or to evaluate the accuracy of model estimates (indicated for each dataset below). Models used are shown in the blue filled boxes and model outputs are listed in the blue open boxes (key estimates reported in detail in the Results are bolded). Connections among the analyses are indicated by the arrows and associated annotations.

## RESULTS

### Epidemic dynamics of the second pandemic wave in NYC

NYC experienced a very large first pandemic wave during spring 2020. Similar to our previous work (*6*), the model-inference system here estimates that 16.6% of the population (95% CrI: 13.6 – 21.5%; or 1.1 – 1.8 million people) had been infected by the end of May 2020 (i.e., end of the first wave; Fig 2D). The city was able to gradually reopen part of its economy during summer 2020 after a 3-month long stay-at-home mandate for all non-essential workers. However, infection resurged beginning in the fall of 2020 and the city experienced a second pandemic wave around Nov 2020 – April 2021 (Fig 2). Following the second wave, an estimated total of 41.7% (95% CrI: 35.4 – 49.3%; or 3.0 – 4.1 million people) had been infected by the end of Apr 2021, including all those infected during the first wave. Note these estimates accounted for under-detection of infections (Fig S1), for which the overall infection-detection rate increased to 37.1% (95% CrI: 33.3 – 43.0%) during the 2^nd^ wave from 15.1% (95% CrI: 11.7 – 18.5%) during the 1^st^ wave. This large number of infections occurred despite the non-pharmaceutical interventions implemented throughout the pandemic and rollout of mass-vaccination starting mid-Dec 2020. In addition, unlike the first wave that predominantly affected older age groups, the second pandemic wave affected all age groups (Figs S2-3).

**Fig 2.**
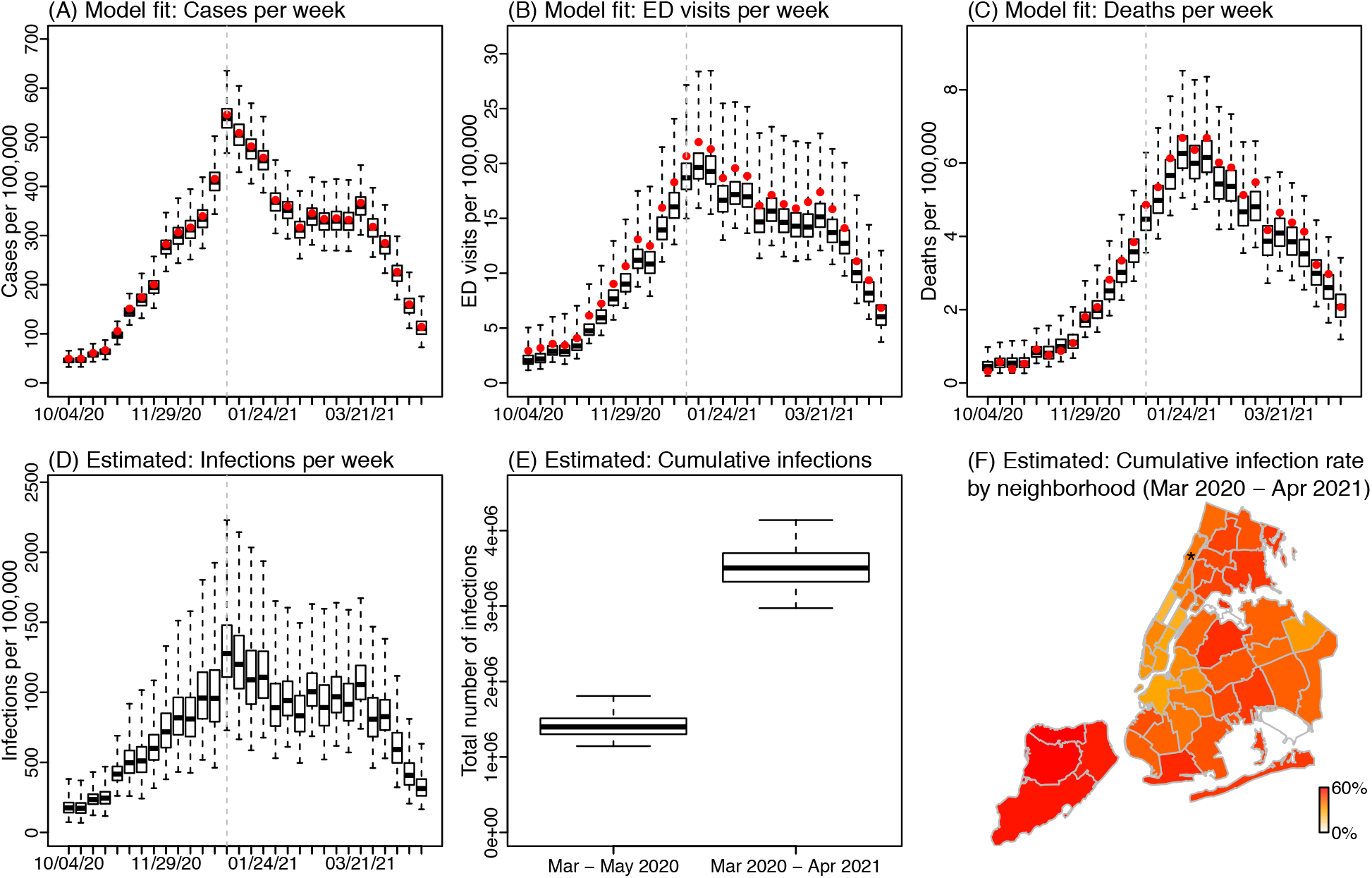
Model fit and key estimates. Upper panel shows model-fit to weekly number of cases (A), ED visits (B), and deaths (C), for all ages combined. Lower panel shows key model-inference estimates of weekly number of infections including those not detected as cases (D), cumulative number of infections in NYC overall (E), and cumulative infection rate by neighborhood (F). Boxes show model estimates (thick horizontal lines and box edges show the median, 25^th^, and 75^th^ percentiles; vertical lines extending from each box show 95% Crl) and red dots show corresponding. For the weekly estimates, week starts (mm/dd/yy) are shown in the x-axis labels. Star (*) in the map indicates the location of the Washington Heights – Inwood neighborhood.

### Transmission rate increased earliest in the neighborhood where B.1.526 was initially identified

The emergence and rapid increase of B.1.526 coincided with the second pandemic wave in NYC. While first reported in Feb 2021 (*2*), testing initially identified the B.1.526 variant in patient samples dated back to early Nov 2020 from the city’s WHts neighborhood (*2*). As such, we first examined potential changes in the transmission rate there. Indeed, prior to the identification of B.1.526, estimated neighborhood relative transmission rate (*b*_*i*_ in Eqn 1; see Methods) in WHts gradually increased, remained at high levels during Nov 2020 – Feb 2021, and decreased to the baseline level afterwards when B.1.526 became a predominant variant citywide (∼40% of all cases sequenced by end of Feb 2021). In comparison, the estimates were relatively stable for other neighborhoods (Fig 3A), suggesting the changes in WHts were likely due to the early spread of B.1.526. Averaging over this period, we estimate that the relative transmission rate in WHts increased by 8.4% (95% CI: -5.8 – 22.5%). Concurrently, the citywide transmission rate increased by 13.3% (95% CI: -21.1 – 47.8%; Fig 3B). These two preliminary estimates in combination suggest that the transmission rate of B.1.526 likely is 22.8% (95% CI: -12.4 – 58.0%) higher than preexisting non-VOC/VOI variants, without accounting for potential change due to immune evasion.

**Fig 3.**
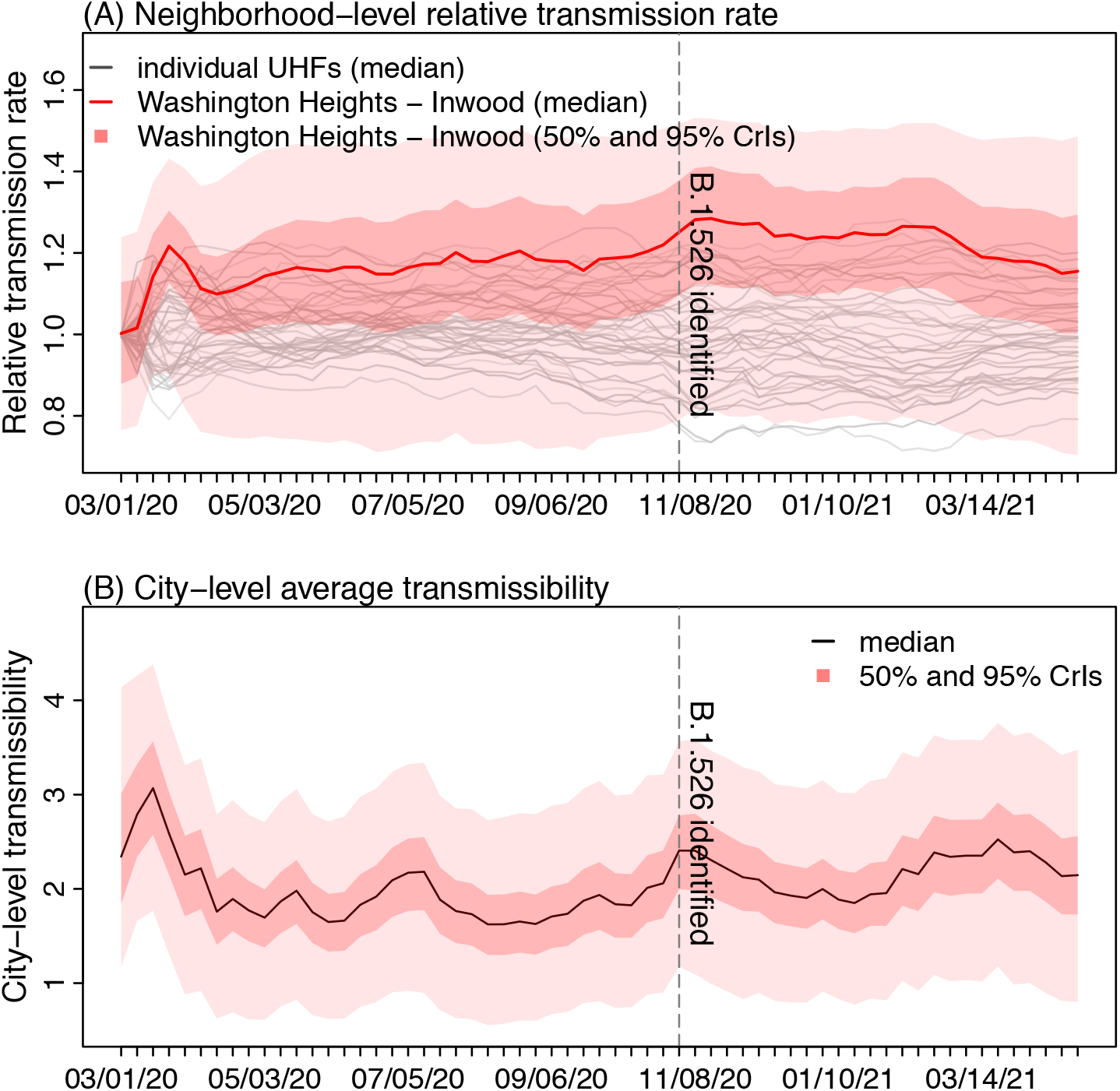
Changes in transmission rate. (A) Changes in neighborhood-level relative transmission rate. (B) Changes in citywide transmission rate. Vertical dashed lines indicate the earliest date B.1.526 was identified as reported in Annavajhala et al. Labels of x-axis show the week starts (mm/dd/yy).

### B.1.526 likely causes a moderate increase in transmissibility (15-25%) and slight immune evasion (0-10%)

We further examine model estimations under a wide range of transmissibility and immune escape settings for B.1.526. Under all three possible scenarios of initial prevalence (i.e., 0.5 – 2.5%, 1.5 – 3.5%, and 0.5 – 3.5%), model simulations consistently show that B.1.526 likely increases transmissibility by 15-30% and can escape immunity in 0-10% of previously infected persons (Fig 4 A-C). Overall, a higher initial prevalence (1.5 – 3.5% at the beginning of Nov 2020) combining with a 15-25% increase in transmissibility and 0-10% immune escape (Fig 4 A-C, middle column; and Fig 4D) generated the most accurate estimates of cases, hospitalizations and deaths as well as variant percentages during the second wave. Model simulations show that, with this moderate increase in transmissibility and small immune escape, B.1.526 was able to outcompete preexisting variants and gradually increase its percentage from Nov 2020 to March 2021; however, afterwards its percentage decreased with the surge of B.1.1.7, a more infectious variant (Fig 4D, bottom right panel).

**Fig 4.**
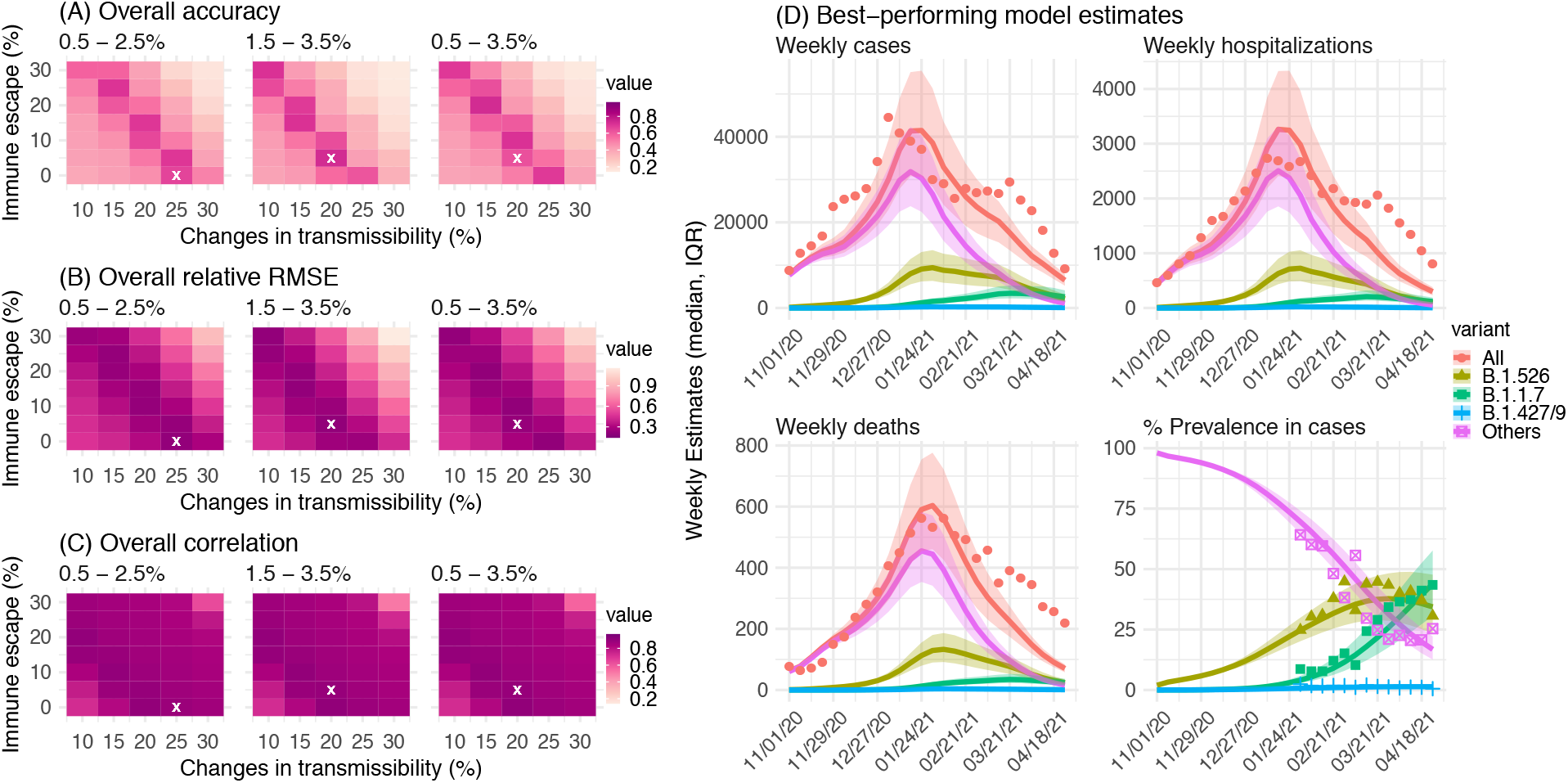
Comparison of different combinations of changes in transmissibility and immune escape property for B.1.526. Left panel shows the overall accuracy (A), relative RMSE (B), and correlation (C) of model estimates under different transmissibility and immune escape settings. White crosses (x) indicate the best-performing parameter combination. Right panel shows model estimates using the overall best-performing parameter combination (i.e., 1.5-3.5% initial prevalence, 15-25% higher transmissibility, and 0-10% immune escape). Lines and surrounding areas show model-simulated median estimates and interquartile range; dots show corresponding observations; colors indicate different variants as specified in the legend. Note that these model simulations used same infection-detection rate, hospitalization-rate and IFR (i.e., average during Nov 2020 – Apr 2021); that is, they did not account for changes in case ascertainment or disease severity by week during this period, due to, e.g., increases in disease severity by the new variants. As such, there were larger deviations from the observations during later months of the simulation with more infections by the new variants.

### B.1.526 likely increases disease severity substantially

During the second wave, estimated IFR increased gradually in later months, particularly among older age groups (Fig 5). During this period (Nov 2020 – Apr 2021), 16-35% of hospital beds and 18-32% of intensive care unit (ICU) beds in NYC were available, suggesting lack of access to healthcare was not a reason behind the IFR increases. In addition, the number of COVID-19-related deaths declined following mass-vaccination in early 2021. Modeling accounting for infections and deaths due to B.1.526, B.1.1.7, and non-VOC/VOI variants suggests that B.1.526 increased IFR in older adults: by 46% (95% CI: 7.4 – 84%) among 45-64 year-olds [absolute IFR: 0.42% (95% CI: 0.31 – 0.54%) vs. 0.29% (95% CI: 0.15 – 0.44%) baseline risk]; 82% (95% CI: 20 – 140%) among 65-74 year-olds [absolute IFR: 1.9% (95% CI: 1.2 – 2.5%) vs. 1.0% (95% CI: 0.57 – 2.5%) baseline risk], and 62% (95% CI: 45 – 80%) among 75+ [absolute IFR: 6.7% (95% CI: 5.9 – 7.4%) vs. 4.1% (95% CI: 2.2 – 6.3%) baseline risk], during Nov 2020 – Apr 2021; overall, B.1.526 increased the IFR by 60% (95% CI: 38 – 82%), compared to estimated baseline risk (Table 1). The analysis restricting to Nov 2020 – Jan 2021 suggests similar IFR increases (Table S3). These estimated IFR increases were lower than for B.1.1.7 but comparable. Of note, the IFRs for B.1.1.7 estimated here were higher than but in line with those reported in the UK [e.g., overall increase: 100% (75-130%) vs. 61% (42–82%) in the UK(*7*)].

**Table 1.** Estimated IFR for different variants and changes compared to the baseline risk estimated for preexisting variants during Oct – Dec 2020, using Eqn 4.

| Age | IFR, baseline (%) | IFR, B.1.526 (%) | IFR, B.1.1.7 (%) | Changes, B.1.526 (%) | Changes, B.1.1.7 (%) | Model goodness-of-fit (R<sup>2</sup>) |
| --- | --- | --- | --- | --- | --- | --- |
| <25 | 0.004 (0.0021, 0.0059) | 0.004 (0.0039, 0.0041) | 0.004 (0.0039, 0.0042) | -0.61 (-3.5, 2.3) | 0.97 (-2.8, 4.8) | 1 |
| 25-44 | 0.04 (0.021, 0.059) | 0.037 (0.034, 0.04) | 0.043 (0.039, 0.047) | -6 (-13, 1.4) | 8.4 (-1.9, 19) | 1 |
| 45-64 | 0.29 (0.15, 0.44) | 0.42 (0.31, 0.54) | 0.51 (0.35, 0.67) | 46 (7.4, 84) | 76 (22, 130) | 0.97 |
| 65-74 | 1 (0.57, 2.5) | 1.9 (1.2, 2.5) | 3.3 (2.4, 4.2) | 82 (20, 140) | 210 (130, 300) | 0.96 |
| 75+ | 4.1 (2.2, 6.3) | 6.7 (5.9, 7.4) | 8 (7, 9) | 62 (45, 80) | 95 (71, 120) | 0.99 |
| all | 0.35 (0.2, 0.58) | 0.56 (0.48, 0.63) | 0.72 (0.61, 0.82) | 60 (38, 82) | 100 (75, 130) | 0.99 |

**Fig 5.**
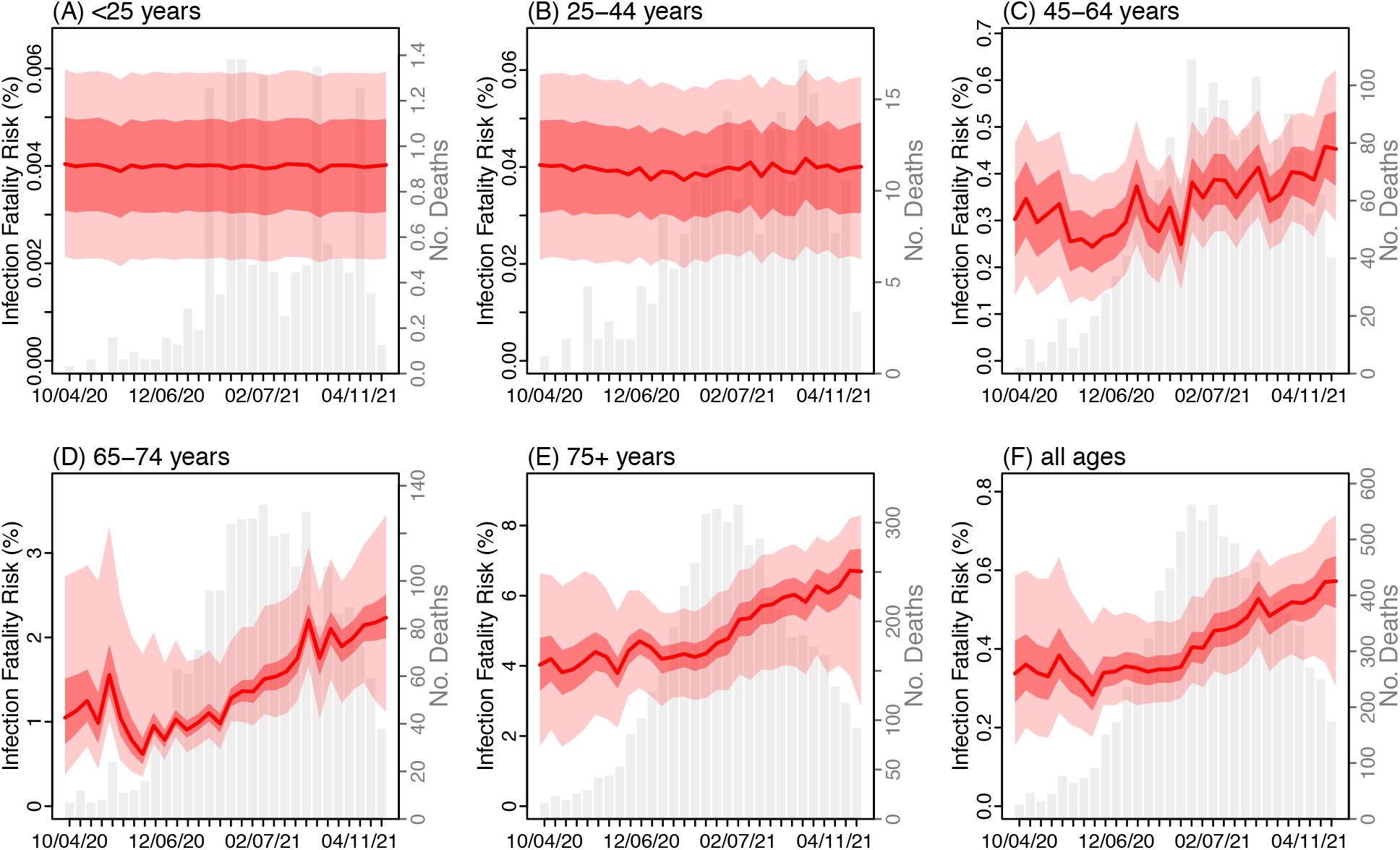
Estimated infection fatality risk. Red lines show the estimated median IFR with surrounding areas indicating the 50% (darker color) and 95% (lighter color) CrI. For comparison, the grey bars show the number of deaths reported for each week from the week of Oct 4, 2020 to Apr 25, 2021. X-axis labels show the week starts (mm/dd/yy).

## DISCUSSION

The B.1.526 variant is one of the SARS-CoV-2 variants designated as a VOI by both the WHO (*1*) and the US CDC (*8*). However, due to a lack of extensive genomic sequencing and contact tracing data particularly during the early phase of its emergence, its key epidemiological properties have not been well characterized. Utilizing multiple epidemiological datasets and comprehensive modeling, here we have estimated the changes in transmissibility, immune escape potential, and disease severity for B.1.526. Results suggest that, compared to preexisting non-VOC/VOI variants, B.1.526 causes a moderate increase in transmissibility and minimal immune evasion; however, it might substantially increase IFR in older adults. As such, continued monitoring of the circulation of this variant is warranted.

Our study offers several lessons for future outbreak response. First, prior to the emergence of B.1.526, the estimated transmission rate in WHts, where it likely emerged, was consistently higher than other neighborhoods in NYC throughout the pandemic. Population characteristics (e.g., household structure) that may contribute to this higher transmission rate need further investigation; however, the higher transmission rate may have facilitated the spread of new mutants between hosts and its emergence population-wide. It is thus important to closely monitor populations with sustained higher transmission rates for new variants, particularly in areas lacking robust and timely sequencing of samples from newly identified cases. In addition, the estimated transmission rate in WHts further increased in conjunction with the emergence of B.1.526; such changes thus may serve as an early indicator for in-depth epidemiological investigation (e.g., to assess changes in circulating variants and transmissibility). A similar approach has been applied in the UK, where subregions with higher estimated growth rates were prospectively investigated, leading to identification of B.1.1.7 as a VOC (*9-12*).

Second, we did not find a higher B.1.526-related IFR among younger age groups (those under 45 years); this finding is consistent with the findings of Thompson et al. (*4*) based on analysis of all sequenced cases, the majority of whom (67%) were under 45 years. However, for older ages, we found substantially higher B.1.526-related IFRs (e.g. >60% higher for those above 65 years). This latter finding appears to be consistent with the report by Annavajhala et al. (*2*) showing resistance of B.1.526 to therapeutic antibodies. Over the course of the pandemic, SARS-CoV-2 IFR has decreased substantially (about a 3-fold difference between the two pandemic waves), likely due to improved medical treatments (e.g., therapeutic antibodies), better patient management, and earlier diagnosis. As older adults are more likely to suffer from severe COVID-19 and thus receive therapeutic antibodies (*13, 14*), the resistance of B.1.526 may render these treatments ineffective despite their prior success against other variants, leading to increases in IFR among older adults. These findings highlight the importance of monitoring the efficacy of therapeutics against different variants and timely update of treatments. In addition, a better understanding of factors contributing to the higher IFRs in certain variants is warranted to inform countermeasures (*15, 16*).

Lastly, our analyses suggest both B.1.526 and B.1.1.7 likely had been spreading in the population for weeks or months prior to detection by the surveillance system (*2, 3, 17*). Expanding genomic sequencing programs for SARS-CoV-2 and improving linkage to epidemiologic data can improve detection of new VOIs/VOCs. Such efforts are underway (e.g., in the US) but more efforts and resources are urgently needed globally. In addition, to support more timely detection and control, targeted screening of key subpopulations (e.g., those prospectively identified from modeling as having high transmission rates) and viral traits (e.g., mutations linked to increased transmissibility and/or immune evasion as done in Annavajhala et al.(*2*)) is needed, as well as timely sharing of key information globally. The documentation of new VOIs/VOCs anywhere in the world then should prompt preparedness measures to detect and rapidly respond to the introduction of those variants into local areas. More fundamentally, to limit emergence of new VOIs/VOCs and end the COVID-19 pandemic, all populations worldwide should have timely access to vaccination, and multiple layers of mitigation efforts are needed until a sufficient portion of the population is protected by vaccination.

Our study also has several limitations. First, most of our analyses are based on population-level data without variant-specific information, given limited variant testing during most of the study period. We circumvented this data deficiency by analyzing estimates of a key subpopulation (e.g., the WHts neighborhood where B.1.526 was initially detected) and leveraging prior knowledge (e.g., estimated IFR prior to B.1.526 emergence). Second, our study did not distinguish the two subclades within the B.1.526 lineage – one containing the E484K mutation and the other containing the S477N mutation. Both the E484K and S477N mutations have been shown to mediate immune escape (*16, 18-20*); in addition, the percentages of these two subclades were similar during our study period, suggesting they likely have similar epidemiological characteristics. Third, while it is likely that the emergence of B.1.526 led to the increase in transmission rate in WHts at the time, we cannot rule out the possibility that other factors contributed to this increase and in turn the emergence of B.1.526. Lastly, there is a likely larger uncertainty in B.1.526-related and B.1.1.7-related IFR estimates for younger ages (those under 45), due to the smaller number of deaths and larger uncertainty in baseline IFR estimates. Future investigation addressing these issues is warranted should a large sample of variant-specific data become available.

In summary, our study has reconstructed the early epidemic trajectory and subsequent rise of B.1.526 in NYC and estimated its key epidemiological properties. Findings highlight the importance of monitoring the viral diversity of SARS-CoV-2, epidemiological characteristics of new variants, and disease severity, as COVID-19 remains a global public health threat.

## MATERIALS AND METHODS

### Study design and data

This study included three interconnected modeling analyses, synthesizing nine epidemiological and population datasets (Fig 1). The first analysis applied a network model-inference system to reconstruct underlying SARS-CoV-2 transmission dynamics in NYC, accounting for under-detection of infection; it also enabled estimation of key population variables and parameters (e.g., the infection rate including those not detected as cases and transmission rate). The second analysis applied a city-level multi-variant, age-structured model to simulate and estimate the changes in transmissibility and immune escape potential for B.1.526 based on the network model-inference estimates and additional data (e.g., variant prevalence data). The last analysis utilized estimates from the first two model systems to estimate variant-specific infection fatality risk (IFR, i.e. the fraction of all persons with SARS-CoV-2 infection who died from the disease), for B.1.526 and B.1.1.7, separately.

For the network model-inference system, we utilized multiple sources of epidemiological data, including confirmed and probable COVID-19 cases, emergency department (ED) visits, and deaths, as well as vaccination data. As done previously (*6*), we aggregated all COVID-19 confirmed and probable cases (*21, 22*) and deaths (*22*) reported to the NYC Department of Health and Mental Hygiene (DOHMH) by age group (<1, 1-4, 5-14, 15-24, 25-44, 45-64, 65-74, and 75+ year-olds), neighborhood of residence (42 United Hospital Fund neighborhoods in NYC(*23*)) and week of occurrence (i.e., week of diagnosis for cases or week of death for decedents). COVID-19-related ED visit data were obtained from the NYC syndromic surveillance system, comprised of all 53 hospital EDs in the city (*24*). This system identified individuals presenting at the EDs with COVID-like-illness (CLI; defined as having a fever and cough or sore throat or respiratory illness, or pneumonia, or a COVID-19 discharge diagnosis code, excluding those with a discharge diagnosis code of influenza only); in addition, CLI patients were matched to electronic laboratory reports of SARS-CoV-2 tests with diagnosis date within ±7 days of ED visit. We estimated the number of COVID-19-related ED visits as the number classified as CLI multiplied by the percentage of those who tested positive for SARS-CoV-2 RNA, stratified by the same age and neighborhood groups in weekly intervals. To account for the impact of vaccination, we also included COVID-19 vaccination data (partially and fully vaccinated, separately), aggregated to the same age/neighborhood strata by week.

In addition, as in our previous study (*6*), we used mobility data from SafeGraph (*25*) to model changes in SARS-CoV-2 transmission rate due to non-pharmaceutical interventions. These data were aggregated to the neighborhood level by week without age stratification.

For the multi-variant model analysis, we additionally utilized four city-level, weekly datasets: 1) COVID-19 confirmed and probable cases, 2) hospitalizations (*26*), 3) deaths, and 4) the percentage of different variants in NYC based on genomic sequencing of samples submitted to the NYC DOHMH Public Health Laboratory and Pandemic Response Laboratory (*4, 27*). The additional hospitalization and variant percentage data were published by the NYC DOHMH (*26, 27*) and accessed on June 22, 2021. We used the variant data from the week starting Jan 31, 2021 to the week starting April 25, 2021 in this analysis, because earlier weeks had very low sample sizes (<200 samples sequenced per week).

This study was classified as public health surveillance and exempt from ethical review and informed consent by the Institutional Review Boards of both Columbia University and NYC DOHMH.

### Network model-inference system

The network model-inference system used here is similar to the approach described in Yang et al.(*6*); however, here we further accounted for waning immunity and vaccination and additionally used COVID-19-related ED visit data for model optimization. Briefly, the model-inference system uses an epidemic model (Eqn 1) to simulate the transmission of SARS-CoV-2 by age group and neighborhood, under implemented public health interventions and mass-vaccination when vaccines became available starting Dec 14, 2020:

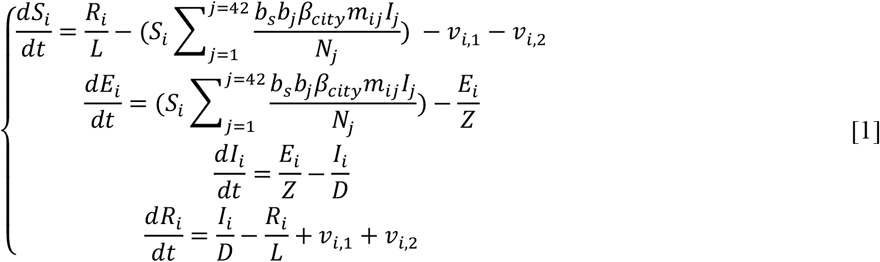

where *S*_*i*_, *E*_*i*_, *I*_*i*_, *R*_*i*_, and *N*_*i*_ are the number of susceptible, exposed (but not yet infectious), infectious, and removed (either recovered, immune, or deceased) individuals and the total population, respectively, from a given age group in neighborhood-*i*. *β*_*city*_ is the average citywide transmission rate; *b*_*s*_ is the estimated seasonal trend (*6*). The term *b*_*i*_ represents the neighborhood-level transmission rate relative to the city average. The term *m*_*ij*_ represents the changes in contact rate in each neighborhood (for *i*=*j*) or spatial transmission from neighborhood-*j* to *i* (for *i*≠*j*) and was computed based on the mobility data (*6*). *Z, D, and L* are the latency period, infectious period, and immunity period, respectively. The term *v*_*i,1*_ represents the number of individuals in neighborhood-*i* successfully immunized after the first dose of the vaccine and is computed using vaccination data and vaccine efficacy (VE) for 1^st^ dose; *v*_*i,2*_ is the additional number of individuals successfully immunized after the second vaccine dose (excluding those successfully immunized after the first dose). Because 97% of vaccine doses administered in NYC during our study period (through April 30, 2021) were the Pfizer-BioNTech or Moderna vaccines, we assumed a VE of 85% fourteen days after the first dose and 95% seven days after the second dose based on clinical trials and real-word data (*28-30*).

Using the model-simulated number of infections occurring each day, we further computed the number of cases, ED visits, and deaths each week to match with the observations (*6*). Similar to the procedure for cases and deaths described in Yang et al.(*6*), to compute the number of ED visits, we multiplied the model-simulated number of new infections per day by the ED-consultation rate (i.e. the fraction of model-simulated persons with new SARS-CoV-2 infections presenting at the EDs), and further distribute these estimates in time per a distribution of time-from-infection-to-ED-consultation (Table S1); we then aggregated the daily lagged, simulated estimates to weekly totals for model inference.

Each week, the system uses the ensemble adjustment Kalman filter (EAKF)(*31*) to compute the posterior estimates of model state variables and parameters based on the model (prior) estimates and observed case, ED visit, and mortality data per Bayes’ rule (*6*). In particular, using this model-inference, we estimated the citywide transmission rate (*β*_*city*_), neighborhood relative transmission rate (*b*_*i*_), and IFR by age group for each week, from the week starting March 1, 2020 (i.e. the beginning of the COVID-19 pandemic in NYC) to the week starting April 25, 2021.

### Multi-variant, age-structured model

Due to model complexity, the model-inference system described above does not account for the circulation of different variants. To model variants, we used a city-level multi-variant, age-structured model (*32*), per Eqn 2:

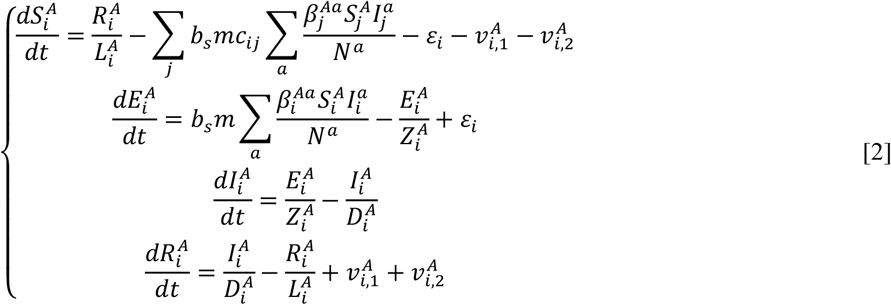

Model variables and parameters in Eqn 2 are similar to those in Eqn 1 with the same symbols. For instance, *β*_*i*_ is the transmission rate for variant-*i*. However, instead of modeling the spatial structure, Eqn 2 focuses on the interactions among different variants (indicated by the subscript, *i*) and age structure (indicated by the superscript, a or A). For age structure, infection in age group *A* by variant-*i* comes from all age groups, per the summation 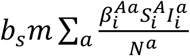 (see the 2^nd^ line in Eqn 2). For variant interactions via cross-immunity, we use a status-based construct similar to Yang et al. (*33*) and Gog and Grenfell (*34*). Specifically, *c*_*ij*_ measures the strength of cross-immunity to variant-*i* conferred by infection of variant-*j* (e.g., close to 0 if it is weak and *c*_*ii*_=1 for infection by the same variant). To compute the depletion of susceptibility to variant-*i* due to infection of variant-*j* (*i*≠*j*), we multiply that infection by *c*_*ij*_; that is, non-specific immunity is scaled by the strength of cross-immunity *c*_*ij*_. As such, the double summation 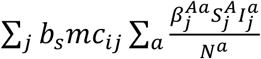 in Line 1 of Eqn 2 represents the depletion of susceptibility due to variant-specific infection (when *i*=*j*) and non-specific infections (for all *i*≠*j*). The vaccination model component 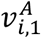 and 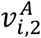 are also variant-specific and can additionally account for the reduction in VE against the new variants if needed; however, here we used the same VE estimates for all variants included (i.e., B.1.526, B.1.1.7, B.1.427 and B.1.429) based on observations (*4, 30, 35, 36*). Additionally, the term *ε*_*i*_ represents travel-related importation of infections of variant-*i* (see Table S2).

We restricted this simulation to Nov 2020 – Apr 2021 (i.e., from the initial identification of B.1.526 to before the further detection and increase of other variants such as Gamma and Delta). In addition to preexisting variants of SARS-CoV-2 prior to Nov 2020 (i.e. “non-VOC/VOI variants” for simplicity) and B.1.526, the model included B.1.1.7 and B.1.427/B.1.429 (combined for simplicity), based on available genomic surveillance data showing consistent detection of these variants during the simulation period and very low levels for others if detected.

Model parameters for B.1.1.7 and B.1.427/B.1.429 were listed in Table S2. For simplicity, we did not account for other VOCs/VOIs variants because their percentages were very low during either analysis period (*27*).

Initial analysis based on the model-inference estimates suggested B.1.526 was around 20% more infectious than non-VOC/VOI variants, without accounting for changes in immunity due to potential immune escape (see details in Results). Therefore, in this analysis, we tested combinations of change in transmissibility ranging from 10 – 30% increases and immune escape ranging from 0 – 30%, both with a 5% increment and ±5% intervals (35 combinations in total). For instance, for the combination centering at 10% transmissibility increase and 0% immune escape, the model is initialized using values in the range of 5-15% (i.e., 10 ± 5%) transmissibility increase and 0-5% (i.e., 0 ± 5% and setting negatives to 0) immune escape. In addition, due to uncertainty on the initial prevalence, we tested three different levels of initial seeding for the week starting Nov 1, 2020, i.e., low (0.5 – 2.5%), high (1.5 – 3.5%), and wider range (0.5 – 3.5%). For reference, Washington Heights – Inwood (WHts), which is the neighborhood where the first patients identified with B.1.526 resided and sought care, constituted 3.2% of the NYC population in 2018. We initialized the model using the model-inference estimates (e.g., population susceptibility and transmission rates by age group; Table S2) and ran the model for each parameter combination 10 times, each with 1000 realizations to account for model stochasticity. Results are summarized from the 10,000 model realizations.

To identify the most plausible combination of transmissibility and immune escape properties for B.1.526, we compared the model-estimated weekly number of cases, hospitalization, and deaths as well as the percentage of the variants to available data. Evaluation was made based on 1) accuracy, i.e., if the observation falls within the model-estimated interquartile range, it is deemed accurate; 2) relative root-mean-square-error (RMSE) between the observed and the model-estimated; and 3) Pearson correlation between the two time-series. Because results show that model accuracy and relative RMSE had a wider spread among the combinations tested (i.e., more distinctive), we first subset those having accuracy within the highest 25^th^ percentile and relative RMSE within the lowest 25^th^ percentile (2-4 out of 35 combinations remained for each setting of initial prevalence); we then selected the one with the highest correlation in the subset as the best-performing and most plausible combination.

### Estimating the changes in IFR due to B.1.526

The network model-inference system enables estimation of the IFR by age group over time. These estimates are made combining all variants and do not distinguish by variant. However, we reasoned that the combined IFR is a weighted average of individual, variant-specific estimates given the relative prevalence of each variant. Accordingly, we built two linear regression models to estimate the variant-specific IFR. Model 1 restricted the analysis to Nov 2020 – Jan 2021 (when the relative prevalence of B.1.1.7 in NYC was likely <10%; n = 14 weeks) and only included two categories of variants:

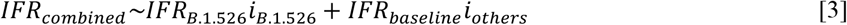

Model 2 extended the analysis to Nov 2020 – Apr 2021 (n = 26 weeks) and included both B.1.526 and B.1.1.7, in addition to other variants:

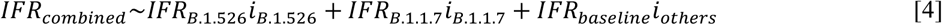

In both models, *IFR*_*combined*_ is the overall IFR for each week, estimated using the model-inference system; *i*_*B*.*1*.*526*_, *i*_*B*.*1*.*1*.*7*,_ *i*_*others*_ are the percentage of infection by the corresponding variant for each week, estimated using the multi-variant age-structured model with the most plausible parameter combination as data are not available. *IFR*_*baseline*_ is the baseline IFR for the preexisting variants, set to the average of model-inference estimates over the period of Oct – Nov 2020 (i.e., prior to the increase of the new variants). The variant-specific IFRs, *IFR*_*B*.*1*.*526*_ and *IFR*_*B*.*1*.*1*.*7*_, are then estimated using the regression models (n = 14 weekly data points for Model 1; and n = 26 weekly data points for Model 2). For either model, the change in IFR due to a given variant is then computed as:

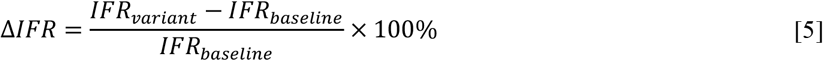

Both model analyses were performed for each age group or all ages combined, separately; we also combined all those aged under 25 as the IFRs were similarly low for the four sub-age groups (i.e. <1, 1-4, 5-14, and 15-24 year-olds).

## Data Availability

The COVID-19 case and mortality data were used with permission under a Data Use and Nondisclosure Agreement between the New York City Department of Health and Mental Hygiene and Columbia University. The New York City Department of Health and Mental Hygiene also has a comprehensive, publicly available data website here:https://github.com/nychealth/coronavirus-data. Additional data sources are detailed in the manuscript. Model code for the multi-variant, age-structured model and a simpler model-inference system using the ensemble adjustment Kalman filter (EAKF; see ref (32)) is available on Zenodo (ref 39).

## Data Availability

The COVID-19 case and mortality data were used with permission under a Data Use and Nondisclosure Agreement between the New York City Department of Health and Mental Hygiene and Columbia University. The New York City Department of Health and Mental Hygiene also has a comprehensive, publicly available data website here: https://github.com/nychealth/coronavirus-data. Additional data sources are detailed in the manuscript. Model code for the multi-variant, age-structured model and a simpler model-inference system using the ensemble adjustment Kalman filter (EAKF; see ref (32)) is available on Zenodo (ref 39).

## Supplemental Tables and Figures

**Table S1.**
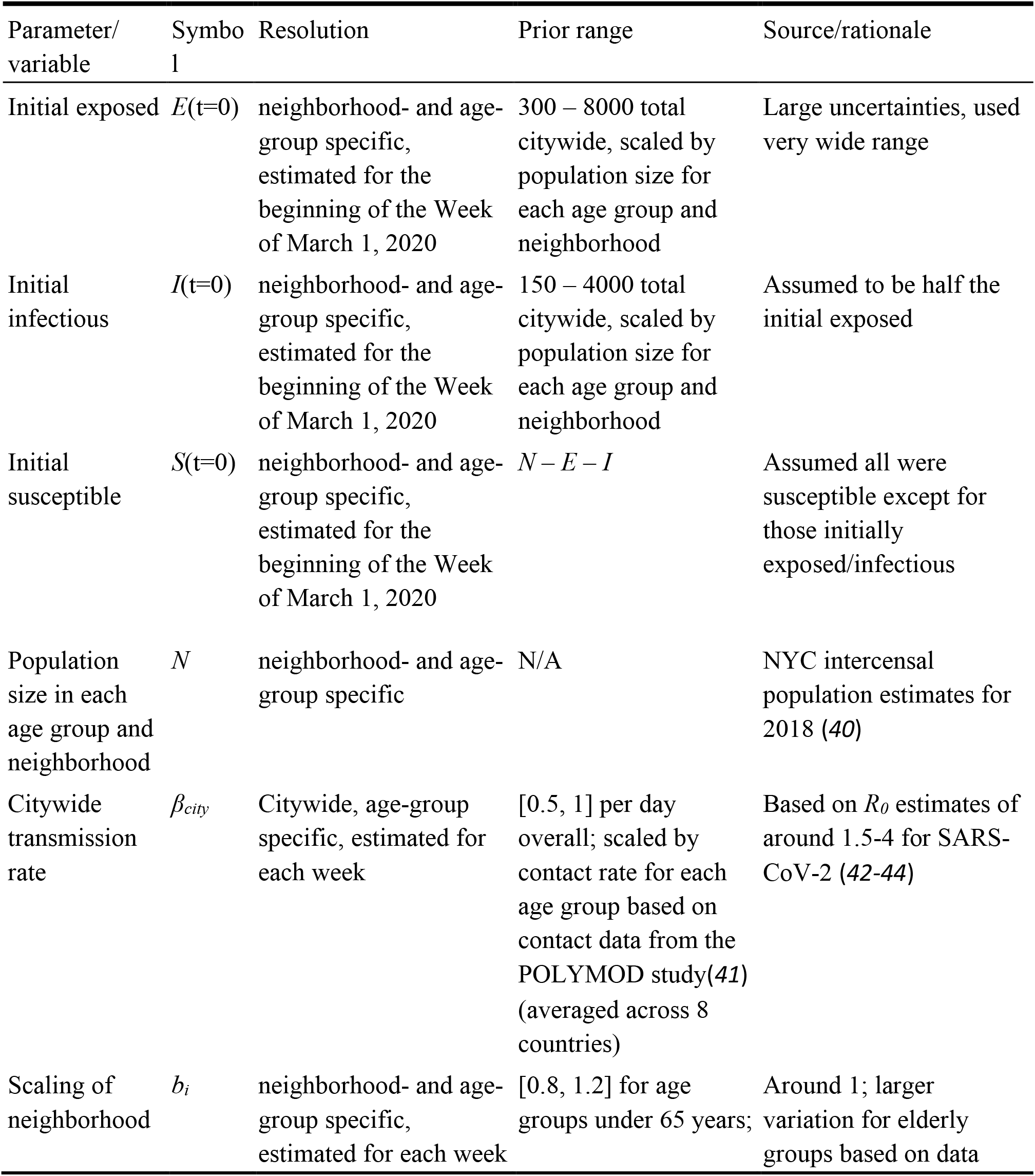
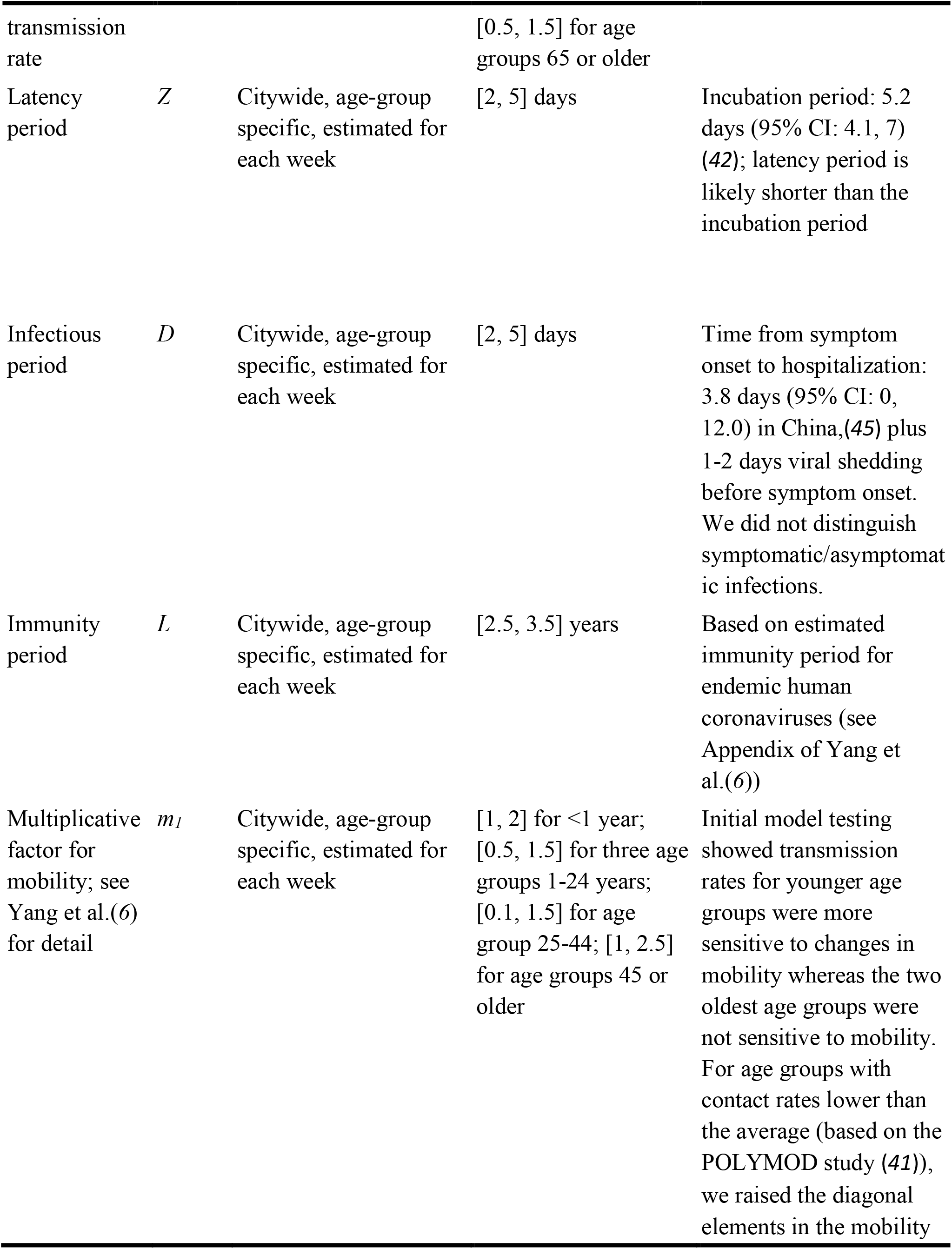
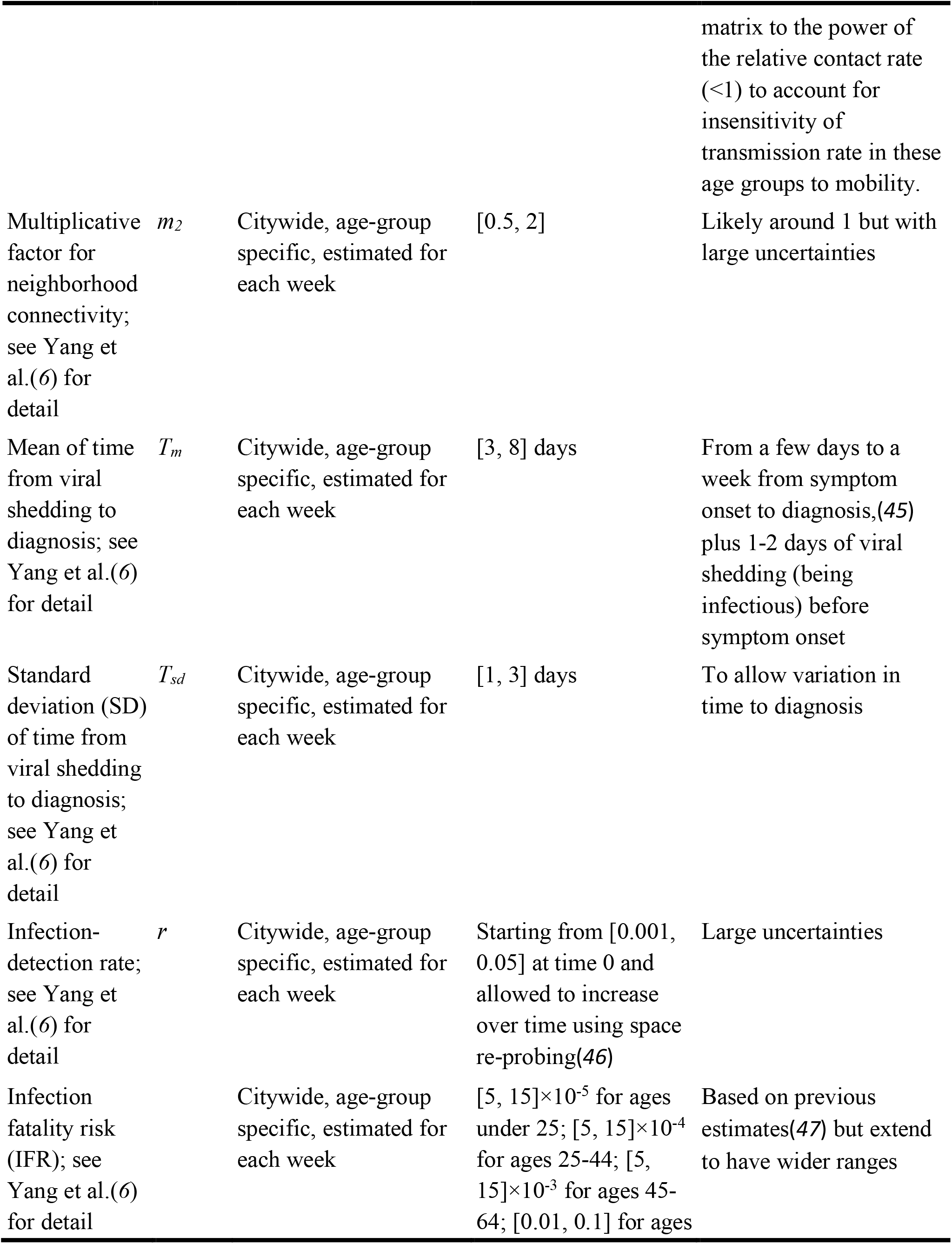
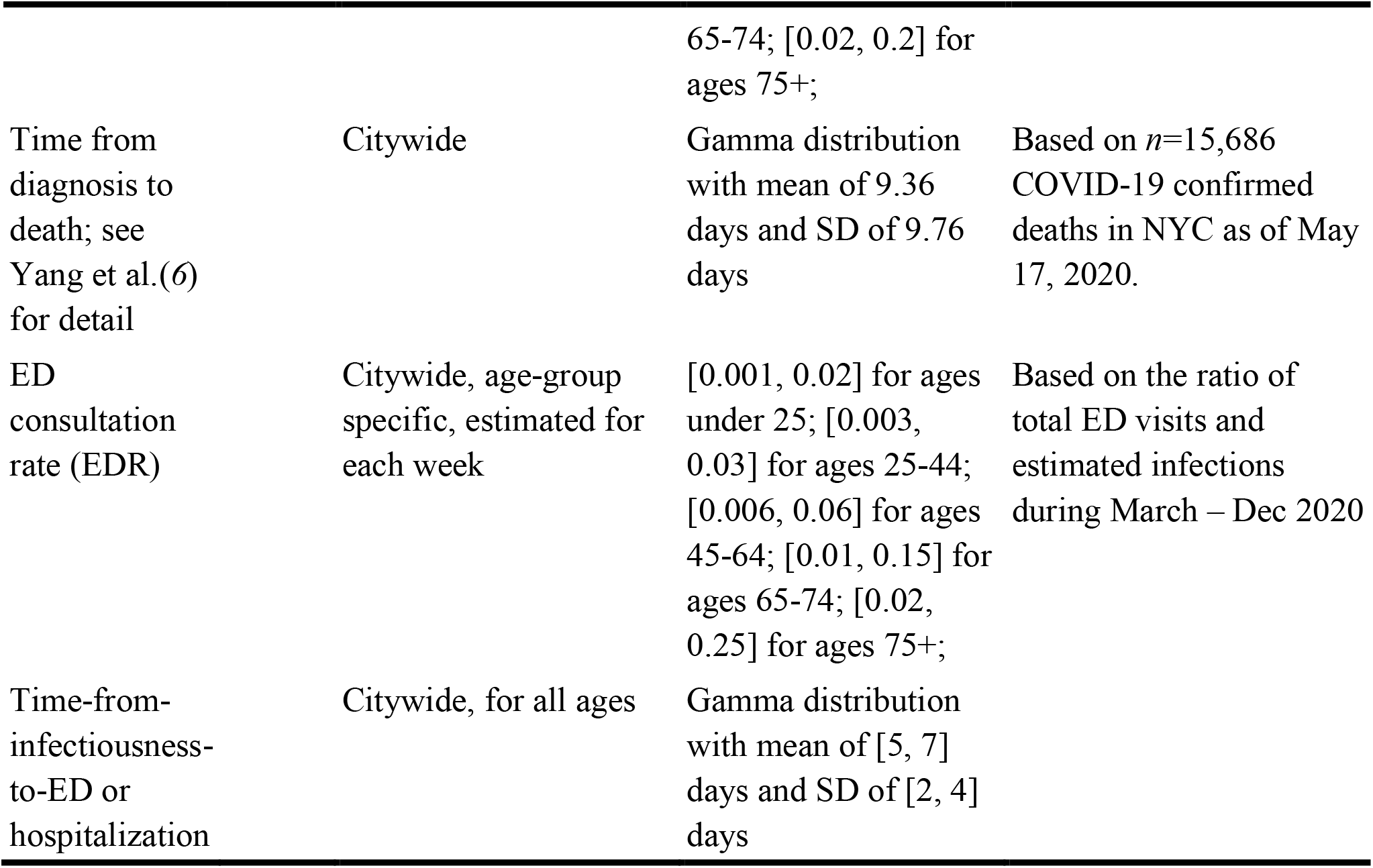
Prior ranges for the network model-inference system. The prior ranges are similar to Table S1 of Yang et al.(*6*) but include additional parameters in Eqn 1. The spatial, temporal, and age resolution of each parameter or variable, estimated in the model-inference system, is specified in the column “Resolution”. Note posterior parameter estimates can extend outside the specified prior ranges.

**Table S2.**
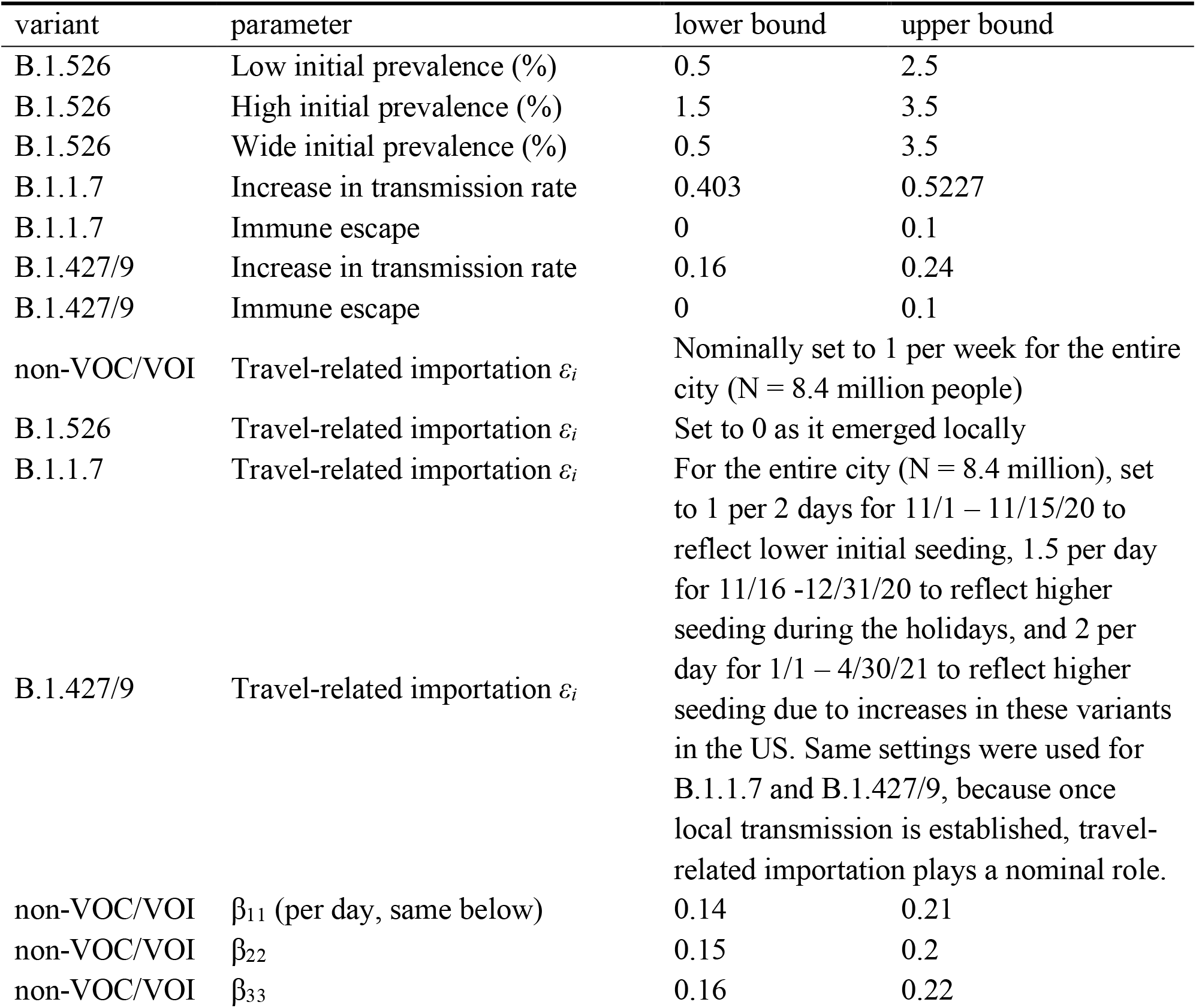
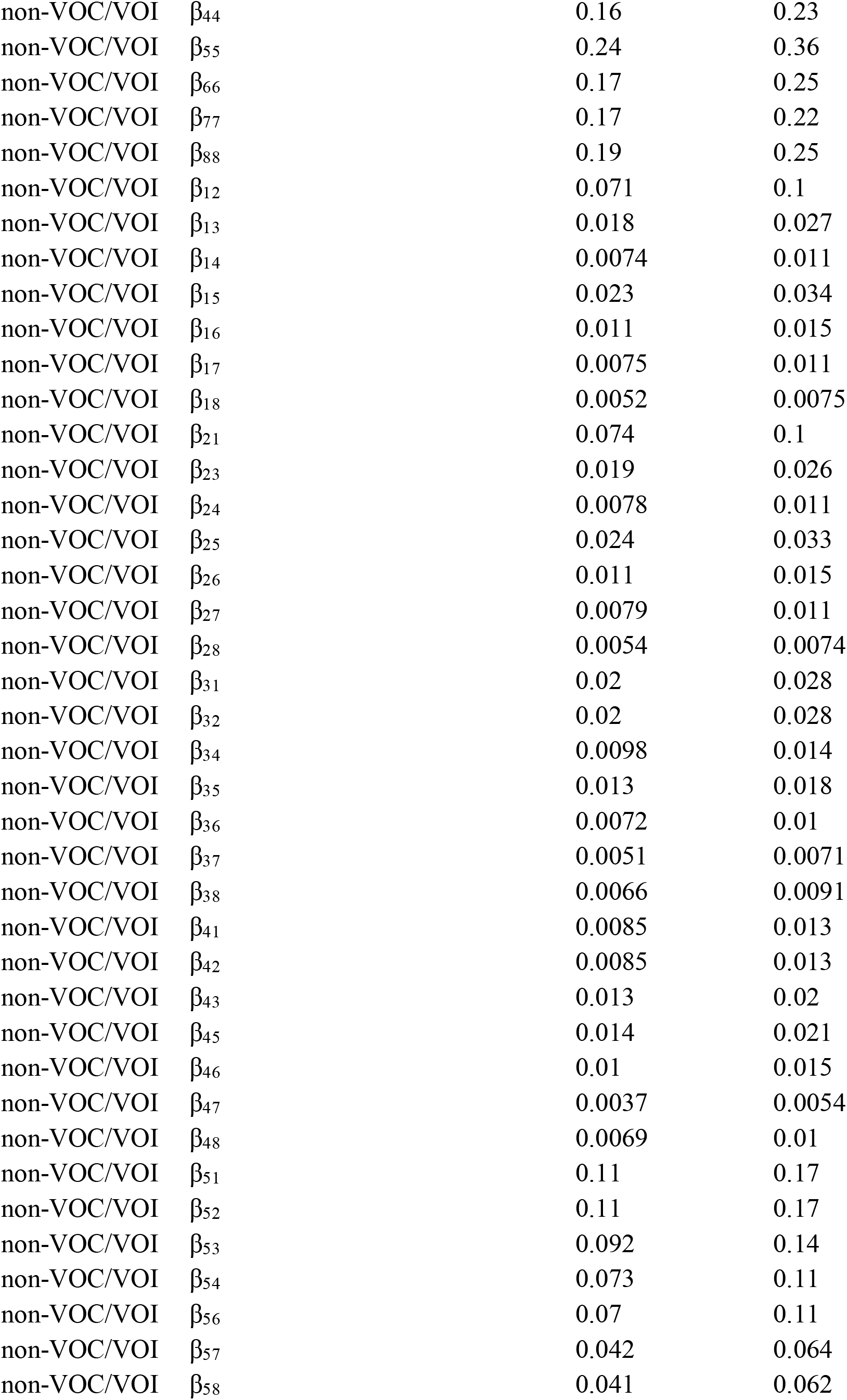
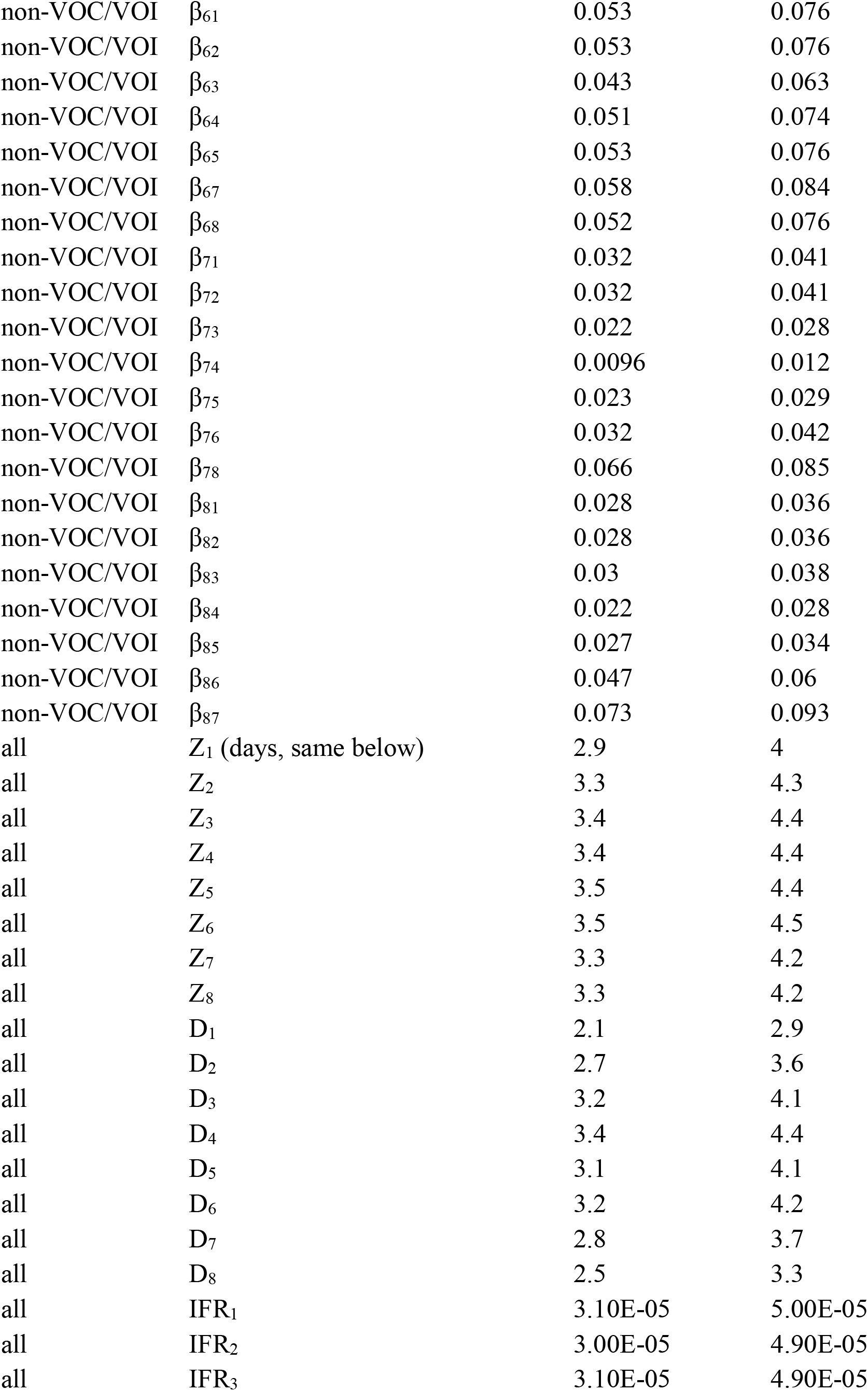
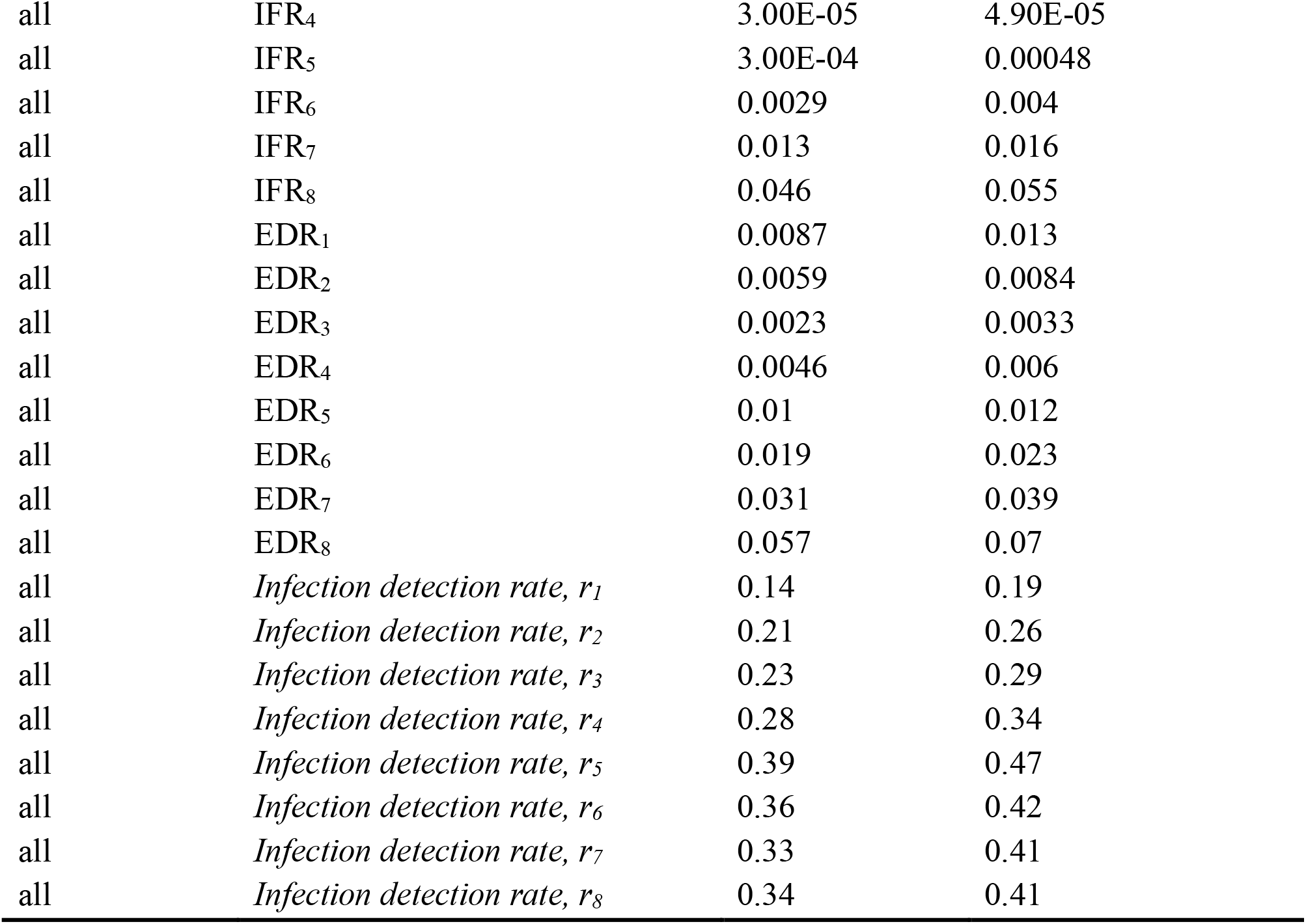
Initial conditions used to simulate co-circulation of different variants in the multi-variant, age-structured model. To partially account for changing infection-detection rate, ED- consultation rate (EDR) and IFR, for these three parameters, we used the model-inference estimates averaged over the entire simulation period (i.e. Nov 2020 – April 2021). For the initial transmission rate (for the preexisting non-VOC/VOI variants), we used the model-inference estimates averaged over the week of 10/25/2020 – the week of 11/7/2020 (i.e. the 3 weeks around the start of simulation). For the rest of model state variables and parameters, we used model-inference estimates made at the week of 10/25/2020. For B.1.1.7, we used the following ranges based on estimates from Yang and Shaman (*32*): 40.3 – 52.3% higher transmissibility (related to estimates for the preexisting non-VOC/VOI variants listed below) and 0 – 10% immune escape; for comparison, contact tracing data from the UK showed that B.1.1.7 was 30- 50% more infectious.(*37*) For B.1.427/ B.1.429, we used the following ranges based on estimates from Deng et al.(*38*): 16 – 24% higher transmissibility and 0-10% immune escape (vs. 21.4 – 27.8% increase in transmission rate in Deng et al.(*38*) without accounting for changes in immunity due to potential immune escape).

**Table S3.** Estimated IFR for different variants and changes compared to the baseline risk estimated for preexisting variants during Oct – Dec 2020, using Eqn 3.

| Age | IFR, baseline (%) | IFR, B.1.526 (%) | Changes,<br>B.1.526 (%) | Model fit,<br>R<sup>2</sup> |
| --- | --- | --- | --- | --- |
| <25 | 0.004 (0.0021, 0.0059) | 0.004 (0.0038, 0.0041) | -0.03 (-3.8, 3.8) | 1 |
| 25-44 | 0.04 (0.021, 0.059) | 0.035 (0.031, 0.038) | -12 (-21, -3.1) | 0.97 |
| 45-64 | 0.29 (0.15, 0.44) | 0.42 (0.24, 0.59) | 43 (-17, 100) | 0.67 |
| 65-74 | 1 (0.57, 2.5) | 1.5 (0.53, 2.5) | 46 (-50, 140) | 0.46 |
| 75+ | 4.1 (2.2, 6.3) | 6.1 (5.1, 7) | 47 (25, 69) | 0.94 |
| all | 0.35 (0.2, 0.58) | 0.5 (0.4, 0.61) | 43 (13, 73) | 0.89 |

**Fig S1.**
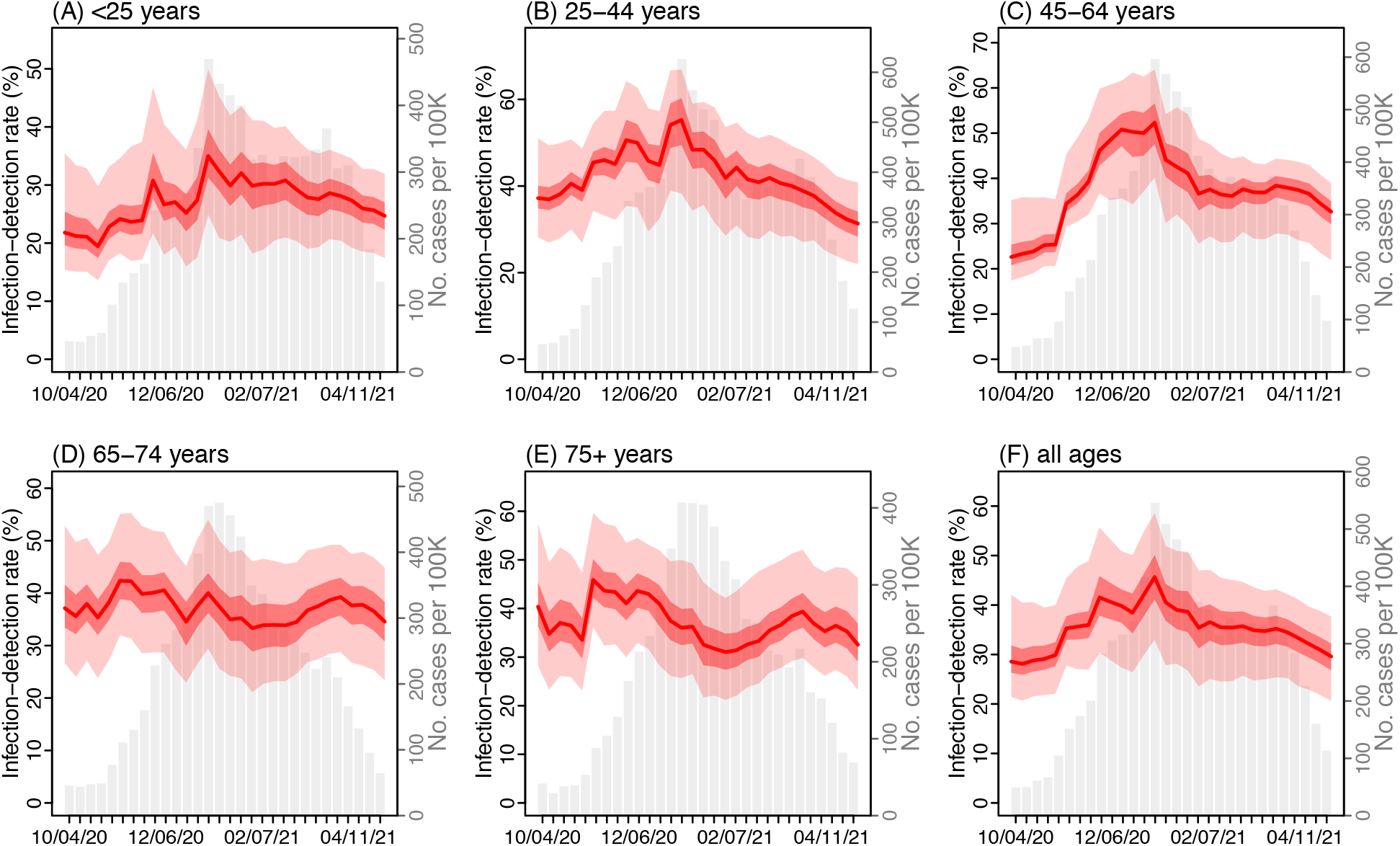
Estimated infection-detection rate by age group. Red lines show the estimated median infection-detection rate with surrounding areas indicating the 50% (darker color) and 95% (lighter color) CrI. For comparison, the grey bars show the number of cases reported for each week from the week of Oct 4, 2020 to Apr 25, 2021. Labels of x-axis show the week starts (mm/dd/yy).

**Fig S2.**
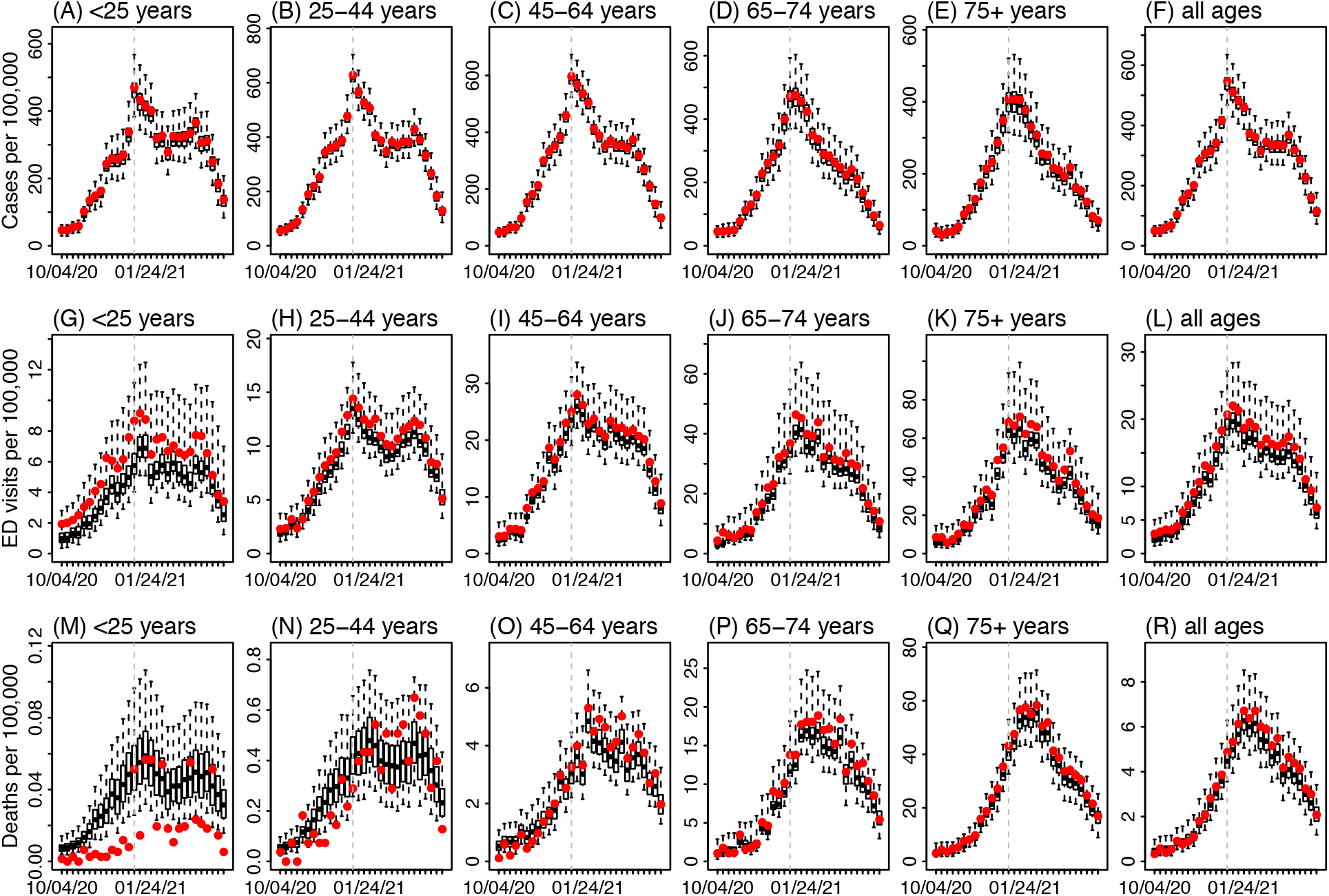
Model-fit by age group. Boxes show model estimates (thick horizontal lines and box edges show the median, 25^th^, and 75^th^ percentiles; vertical lines extending from each box show 95% Crl) and red dots show corresponding.

**Fig S3.**
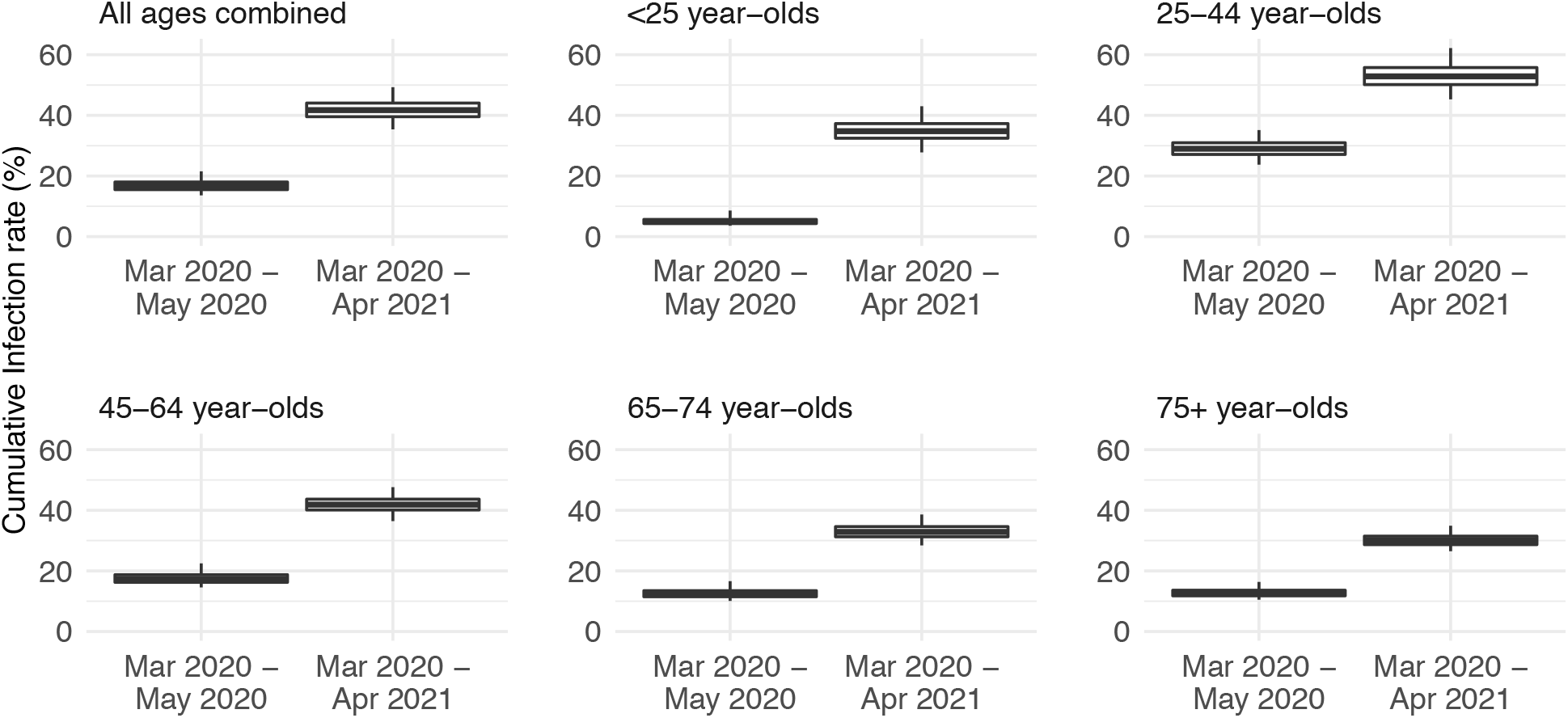
Estimated cumulative infection rates by age group. Thick horizontal lines and box edges show the median, 25^th^, and 75^th^ percentiles; vertical lines extending from each box show 95% Crl.

## Acknowledgments

We thank Columbia University Mailman School of Public Health for high performance computing, Safe Graph (safegraph.com) for providing the mobility data, and Sasikiran Kandula at Columbia University for compiling the mobility data used in this study. We thank the NYC DOHMH Incident Command System Surveillance and Epidemiology Section for processing, cleaning, and managing COVID-19 surveillance data, the NYC DOHMH Public Health Laboratory and Pandemic Response Laboratory for generating and analyzing sequence data, and Iris Cheng, Mohammed Almashhadani, Charles Ko, and Jaimie Shaff from the NYC DOHMH for providing the vaccination data and helpful suggestions on the manuscript.

## Author contributions

WY designed the study, conducted the analysis, and wrote the first draft; SKG contributed to study coordination and specification of COVID-19 case data, and provided input on parameter estimation; ERP led aggregation and provision of COVID-19 case data; LG contributed to management of COVID-19 case data; WL provided the COVID-19-associated mortality data; RM and RL oversaw the collection of and provided the COVID-19 ED data; SH and JW provided input on SARS-CoV-2 variants and interpretation of the NYC variant percentage data; AF oversaw data collection and management processes at DOHMH. All authors contributed to the final draft.

## Data and materials availability

The COVID-19 case and mortality data were used with permission under a Data Use and Nondisclosure Agreement between the New York City Department of Health and Mental Hygiene and Columbia University. The New York City Department of Health and Mental Hygiene also has a comprehensive, publicly available data website here: https://github.com/nychealth/coronavirus-data. Additional data sources are detailed in the manuscript. Model code for the multi-variant, age-structured model and a simpler model-inference system using the ensemble adjustment Kalman filter (EAKF; see ref (*32*)) is available on Zenodo (*39*).

